# Patients with Alzheimer’s disease have increased cellular amyloid uptake

**DOI:** 10.1101/2022.01.12.22269196

**Authors:** Dmitry V. Zaretsky, Maria V. Zaretskaia, Yaroslav I. Molkov, for the Alzheimer’s Disease Neuroimaging Initiative

**Author notes:** Data used in the preparation of this article were obtained from the Alzheimer’s Disease Neuroimaging Initiative (ADNI) database (adni.loni.usc.edu). As such, the investigators within the ADNI contributed to the design and implementation of ADNI and/or provided data but did not participate in analysis or writing of this report. A complete listing of ADNI investigators can be found at: http://adni.loni.usc.edu/wp-content/uploads/how_to_apply/ADNI_Acknowledgement_List.pdf.

## Abstract

Amyloid plaques are the main signature of Alzheimer’s disease (AD). Beta-amyloid (Aβ) concentration in cerebrospinal fluid (CSF-Aβ) and the density of amyloid depositions have a strong negative correlation. However, AD patients have lower CSF-Aβ levels compared to cognitively normal people even after accounting for this correlation. The goal of this study was to infer variations of parameters in Aβ metabolism of AD patients that underlie this difference using data from the Alzheimer’s Disease Neuroimaging Initiative cohort.

We found that AD patients had dramatically increased rates of cellular amyloid uptake compared to individuals with normal cognition (NC). A group with late-onset mild cognitive impairment (LMCI) also exhibited stronger amyloid uptake, however this was less pronounced than in the AD group. Estimated parameters in the early-onset MCI group did not differ significantly from those in the NC group.

Aβ cytotoxicity depends on both the amount of peptide internalized by cells and its intracellular degradation into toxic products. Based on our results, we speculate that AD and LMCI are associated with increased cellular amyloid uptake which leads to faster disease progression, whereas the early-onset MCI may be mediated by the increased production of toxic amyloid metabolites.

## INTRODUCTION

### Biomarkers of beta-amyloid metabolism *in vivo* and their relevance to Alzheimer’s disease

Dr. Alois Alzheimer described a certain kind of dementia (later named after him [13]) as having specific histopathological correlates - senile plaques and neurofibrillary tangles [1]. Since then, it has been quite common to assume that the etiology and pathogenesis of this disease is associated with the main component of the extracellular deposits - beta-amyloid protein (Aβ). Therefore, many clinical studies concentrated on the possibility of using available measures of Aβ presence for diagnostic purposes and predicting disease progression.

For most of the twentieth century, the brain could only be studied *postmortem*. With the increased sensitivity of analytical methods, measuring the concentration of various Aβ peptides (including 42 amino acids long peptide Aβ42) in cerebrospinal fluid (CSF) became possible [5, 25]. Then, positron emission tomography (PET)-measurable labels such as FDA-approved florbetapir and florbetaben, which bind to and accumulate in the senile plaques, became available [22]. Patients with cognitive decline due to Alzheimer’s disease have significantly more amyloid deposits. The appearance of deposits also has a prognostic value as patients with amyloid deposits appear much more prone to decline cognitively compared to amyloid-negative patients [30].

These two indices (the concentration of beta-amyloid in CSF and the density of amyloid deposits) not only have a strong diagnostic potential but are also interconnected. High levels of amyloid deposits are associated with significantly decreased Aβ42 levels in the CSF [11, 21, 39]. Due to the correlation between these two parameters, the overall diagnostic accuracy of CSF-Aβ42 and PET studies appears similar with PET having higher specificity [22]. Despite their significant correlation, CSF-Aβ42 levels and PET imaging have independent predictive powers for different Alzheimer’s disease markers, such as AD diagnosis itself, hippocampal volume, and brain metabolism [21].

Based on analysis of the data from the Alzheimer’s Disease Neuroimaging Initiative (ADNI), Sturchio et al [37] reported that normal cognition in patients with brain amyloidosis is associated with higher CSF-Aβ42. In their study, only amyloid PET-positive participants from the ADNI database, for whom a neuropsychological evaluation was performed and a CSF specimen was collected within 1 year from a PET scan, were selected for analysis. The levels of soluble Aβ42 were statistically lower in the following order: (Alzheimer’s disease, AD) < (mild cognitive impairment, MCI) < (normal cognition, NC). The same phenomenon was present in each group when amyloid-positive patients were separated into three tertiles – with high, medium and low amyloid load; patients with normal cognition had higher CSF-Aβ42 compared with patients with AD [37].

This observation can be deduced from and, thus, confirmed by the data of Mattsson et al. [21]. In their study, all available patients from the ADNI dataset were used. The correlation between CSF-Aβ42 and florbetapir PET with an AD diagnosis were estimated as the regression coefficients and compared between models, which considered the effect of each index either individually or combined. The model that used both indices as independent variables provided a statistically significantly better fit than models that considered either of the indices alone. The combination of CSF-Aβ42 and PET also provided a better prediction for cognitive assessment than the individual indices [21] as CSF-Aβ42 correlated with cognitive status in individuals with the same amyloid load. In other words, for a given amyloid load, cognition indices correlate with CSF-Aβ42 levels. This correlation was also observed in patients with a positive amyloid load (the subset analyzed in the study by Sturchio et al. [37]).

Thus, CSF-Aβ42 concentration and beta-amyloid deposits are partially independent correlates of cognitive function [21]. More specifically, unlike PET which positively correlates with AD diagnosis, the correlation between CSF-Aβ42 and an AD diagnosis or cognitive assessment is negative. Thereby, Mattsson et al. [21] essentially demonstrated that normal cognition is associated with higher CSF-Aβ42 levels, however, they did not specifically emphasize this fact and did not go further than stating that CSF-Aβ42 levels provide additional information independent of amyloid load measured by PET. In contrast, Sturchio et al. [37] attempted to interpret this fact by speculating that soluble amyloid has an unknown “good” function, so that insufficient soluble amyloid may be the reason for neuronal loss (as opposed to the idea that exceedingly high levels of amyloid deposits are “bad”). While this speculation sounds logical, it cannot explain why CSF-Aβ42 levels in patients with normal cognition but high amyloid load can be the same as CSF-Aβ42 levels in patients with AD and low amyloid load [37].

To interpret the interaction between the two major biomarkers of AD, we estimated the parameters of beta-amyloid turnover by fitting a mathematical model to the data from the ADNI dataset.

## METHODS

### Clinical dataset

We used non-personalized data which were obtained from the Alzheimer’s Disease Neuroimaging Initiative (ADNI) (http://adni.loni.usc.edu/). The ADNI was launched in 2003 as a public-private partnership, led by Principal Investigator Michael W. Weiner, MD. The primary goal of ADNI has been to test whether serial magnetic resonance imaging, positron emission tomography (PET), other biological markers, and clinical and neuropsychological assessment can be combined to measure the progression of mild cognitive impairment (MCI) and Alzheimer’s disease (AD). The study protocol for ADNI was approved by the local ethical committees of all participating institutions and all participants signed informed consent.

Our analysis included all ADNI participants for whom the ascertainment of normal cognition (NC), MCI, or AD, as well as a CSF collection, were made within one year from a PET scan identifying brain amyloidosis. The number of research subjects in the AD, NC, late-onset MCI (LMCI), and early-onset MCI (EMCI) groups was 143, 416, 340, and 476, respectively. All subjects were evaluated between June 2010 and February 2019. Amyloid positivity was defined by PET data according to ADNI guidelines as a standard uptake value ratio (SUVR) at or above 1.08 for [18]F-florbetaben or 1.11 for [18]F-florbetapir, with a higher SUVR indicating a greater amyloid plaque burden. Details regarding PET acquisition are described in previous publications and on the ADNI website (www.adni-info.org). Given the use of two different amyloid PET-tracers, SUVR levels were converted to centiloids (CL) using the specific equation for each tracer as provided by ADNI.

### The effect of CSF-Aβ42 levels on the fraction of subjects with AD in subpopulations with low and high densities of amyloid deposits

This part of the study included only AD patients and NC subjects who had an amyloid load exceeding 20 CL. All research subjects in this study were divided into two groups: those with a low load (20<CL<60) and those with a high load (CL≥60).

Within each group, we calculated the fraction of patients with an AD diagnosis as a function of CSF-Aβ42. To do this, we built a histogram using bins with widths of 500 pg/ml and left borders increasing from 50 pg/ml in increments of 50 pg/ml. The data for each patient could appear in multiple adjacent bins, and only bins that had more than 10 study subjects were used. We calculated the average CSF-Aβ42 and the fraction of AD patients in each bin. With the assumption that the fraction follows a binomial distribution, the standard deviation of the fraction of AD patients in each bin was calculated as 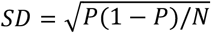, where *P* is the fraction and *N* is the total number of research subjects in the bin. Sigmoidal approximation was performed using the Solver module in MS Excel.

### The single compartment model of cerebral amyloid turnover

The concentration of soluble amyloid in the interstitial fluid (ISF), which we denote as [ISF], is defined by several processes: 1) synthesis by cells, 2) filtration of the protein into the CSF, 3) aggregation into non-soluble plaques, and 4) uptake by cells (see Fig. 2). The model is based on several assumptions:

1. Synthesis rate 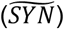 is independent of both interstitial Aβ42 and the density of plaques.
2. The rate of removal of the protein through the CSF is a product of the CSF removal rate (*FLOW*_*CSF*_) and CSF-Aβ42 ([CSF]): *FLOW*_*CSF*_ · [*CSF*].
3. The concentrations of the soluble beta-amyloid in the ISF and the CSF have a similar order of magnitude and are correlated [7, 8]. The model assumes a linear relationship between the concentrations of soluble Aβ42 in the ISF and the CSF with a coefficient of transfer *K*_*T*_: [*CSF*] = *K*_*T*_ · [*ISF*].
4. Existing plaques serve as seeds for aggregation of soluble Aβ42 in the ISF. The rate of loss of soluble Aβ42 in the ISF due to aggregation is the product Aβ42 concentration in the ISF, the concentration of plaques ([PET], calculated from the intensity of the PET signal), and the coefficient of aggregation *K*_*a*_: *K*_*a*_ · [*PET*] · [*ISF*].
5. The rate of cellular uptake of soluble Aβ42 is proportional to the interstitial concentration [ISF] with a coefficient of uptake *K*_*u*_: *K*_*u*_ · [*ISF*].

We assume that at any given moment, [ISF] is in equilibrium:

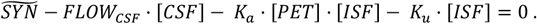

Substituting [*ISF*] with [*CSF*]/*K*_*T*_,

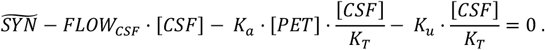

Rearranging this equation, [CSF] can be expressed as a function of [PET]:

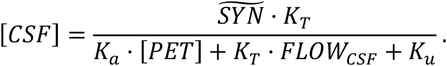

Using the normalized parameters 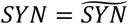 · *K*_*T*_/*K*_*a*_ and *KF* = (*K*_*T*_ · *FLOW*_*CSF*_ + *K*_*u*_)/*K*_*a*_, the equation becomes

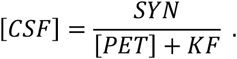

The equation has two parameters, *SYN* and *KF*, which will be referred to as the amyloid synthesis rate and the amyloid removal rate, respectively, in the rest of this manuscript. It should be noted that the normalized removal rate *KF* is a sum of the normalized amyloid removal rate through the CSF (*FLOW*_*CSF*_ · *K*_*T*_/*K*_*a*_) and the normalized cellular amyloid uptake rate (*K*_*u*_/*K*_*a*_).

### Parameter inference

To infer the values of the parameters SYN and KF for each of the four groups (NC, EMCI, LMCI, and AD), we used the CSF-Aβ42 and the composite amyloid density of each research subject (based on data provided by the ADNI). We assumed that the conditional probability distribution *p*([*CSF*]|[*PET*]) is log-normal. Therefore, the posterior probability density function (PDF) of the parameters can be represented as

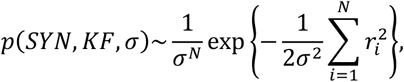

where the residual *r*_*i*_ = ln[*CSF*]_*i*_ - ln(*SYN*/([*PET*]_*i*_ + *KF*)), *σ* is an unknown standard deviation, *N* is the number of participants in the group, and ln(.) is the natural log function. To calculate the marginal PDF for *SYN* and *KF*, we integrated the posterior PDF over *σ* as follows.

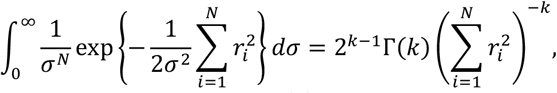

where *k* = (*N* − 1)/2 and Γ(*k*) is the gamma function. Therefore, the following holds.

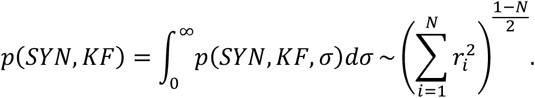

This marginal parameter distribution was used to calculate the confidence regions on the (*SYN, KF*)-plane as well as the standard errors for individual parameters.

### Calculations and statistical comparisons

Calculations were performed using MS Excel and custom written C++ programs. Comparison of parameter values between groups was performed using a z-test, where significance was defined as p<0.05. Values are presented as mean ± SEM.

## RESULTS

### Patients with higher CSF Aβ42 levels are less prone to AD

The probability of an AD diagnosis in randomly selected research subjects from the study cohort clearly correlates with their CSF-Aβ42 levels. To illustrate this, we calculated the fraction of AD patients as a function of their CSF-Aβ42 in two groups both composed of the AD and NC participants (Fig. 1). The first group included participants with a low amyloid load (see Methods), and the second group included participants with a high amyloid load. The dependence appeared much more pronounced in research subjects with a high amyloid load as the fraction of AD patients increased from 10% at the highest observed CSF-Aβ42 to 65% at the lowest observed CSF-Aβ42 values. Due to a small number of observations, we could not reliably calculate the percentage of the patients with AD diagnosis at the highest CSF-Aβ42 values, so we used a sigmoid approximation of the curve for extrapolation and found that the fraction of AD patients approached zero in subjects with both high and low amyloid load as CSF-Aβ42 exceeds 1 ng/ml. The maximal percentage of patients with AD diagnosis was observed when CSF-Aβ42 is the lowest. We estimated it to be 27% in subjects with low amyloid deposit density and 65% in those with high deposit density.

**Figure 1.**
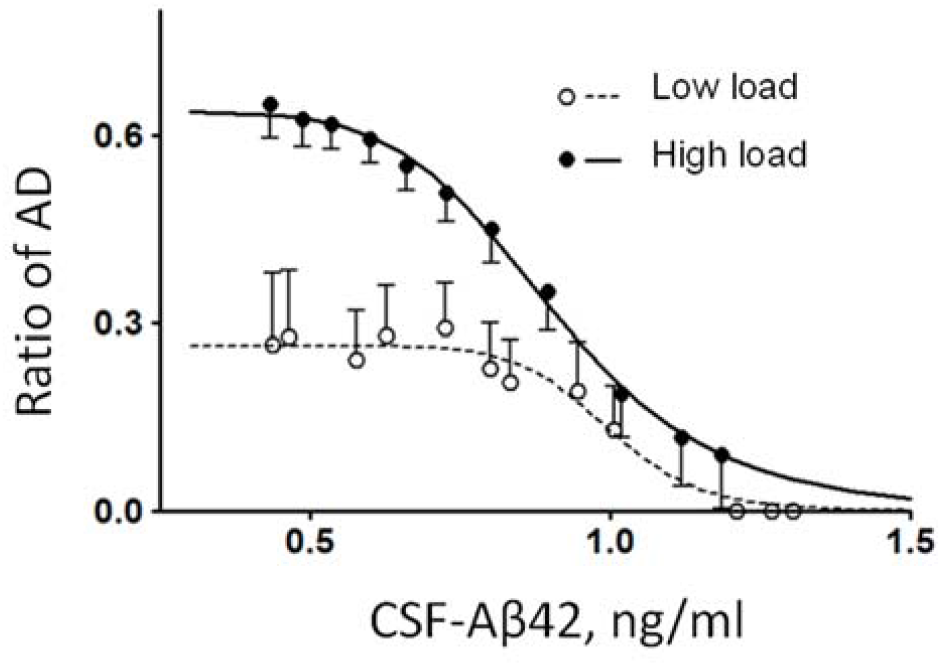
The relative frequency of AD patients as a function of the concentration of CSF-Aβ42 in subpopulations with low and high amyloid deposition density (amyloid load). Error bars indicate standard deviations. Lines represent the best fits by a sigmoid function.

### The relationship between CSF-Aβ42 levels and amyloid load can be described using a simple mathematical model

We used a single-compartment model to describe the beta-amyloid concentration in the interstitial fluid. The model considers Aβ synthesis by cells, its removal by CSF, cellular uptake by endocytosis, and aggregation into amyloid deposits (Fig. 2, see details in Methods). We found that the steady state of free amyloid concentration corresponds to a specific relationship between CSF-Aβ42 (denoted by [*CSF*]) and the density of amyloid deposits ([*PET*]):

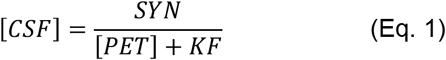

where *SYN* is a parameter representing the synthesis rate and *KF* is a sum of the rate of amyloid removal through the CSF and the cellular amyloid uptake rate. Both *SYN* and *KF* are normalized (see Methods for details). Equation (1) readily explains the negative correlation between CSF-Aβ42 and the density of amyloid deposits (for graph, see [10]).

**Figure 2.**
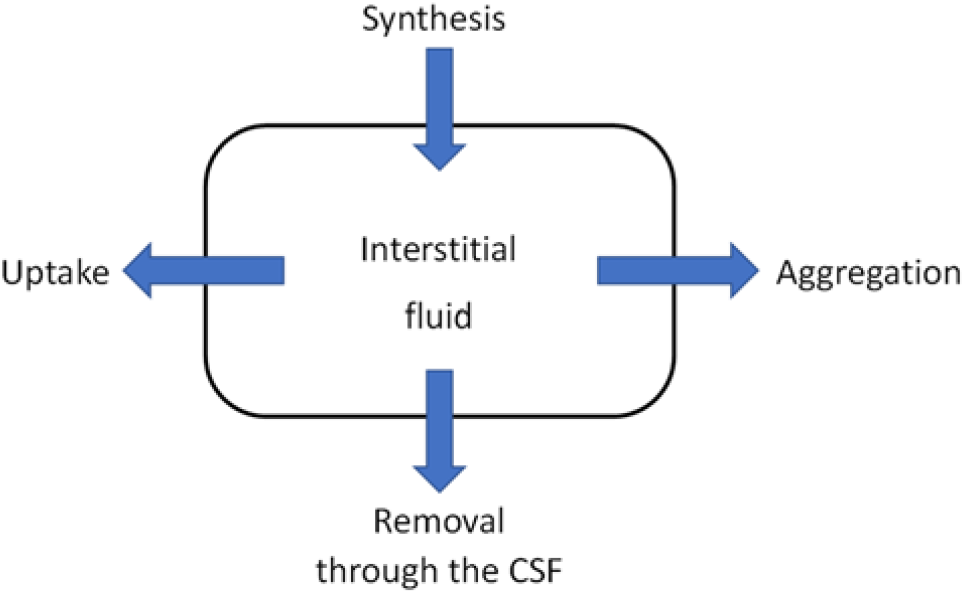
A schematic of the single compartment model of beta-amyloid turnover used to describe the mathematical relationship between CSF-Aβ42 and amyloid load in the brain.

We then used the ADNI data set to infer unknown values of the parameters by fitting the model to the data available for groups with different diagnoses under an assumption that CSF-Aβ42 is measured as a log-normally distributed quantity with unknown variance. Therefore, the statistical inference was performed based on the marginal probability density function (PDF) for *SYN* and *KF* obtained by integration of the posterior PDF over the error variance (see Methods).

### AD patients have higher amyloid removal rate compared to subjects with normal cognition

Fig. 3A shows data on CSF-Aβ42 vs. PET-measured amyloid load in subjects with normal cognition (NC) as well as in patients with late-onset mild cognitive impairment (LMCI) or an AD diagnosis. The major difference between the clouds corresponding to the different groups is that AD patients tend to have greater amyloid loads compared to NC subjects who do show a lesser accumulation of amyloid deposits on average. Patients with LMCI are more broadly distributed as this group includes both individuals with significant amyloid deposition (comparable to AD patients) as well as those who have a low amyloid accumulation (similar to a majority of NC subjects). As far as CSF-Aβ42 levels are concerned, the shape of the cloud is very similar for all three groups, and any differences in the relationship between CSF-Aβ42 and amyloid load are hardly distinguishable to the naked eye.

**Figure 3.**
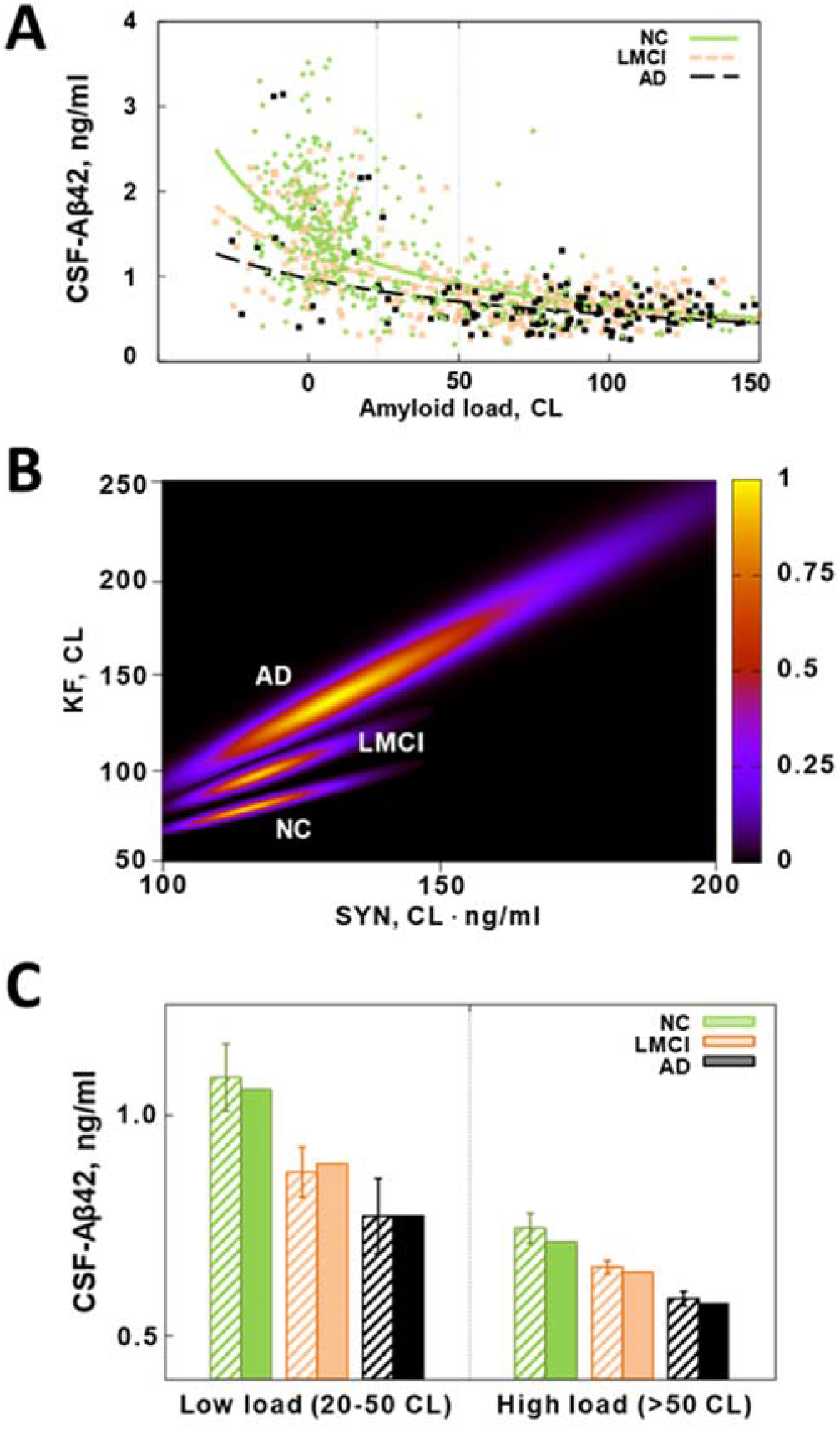
A comparison of beta-amyloid turnover parameters in subjects with normal cognition (NC), patients with Alzheimer’s disease (AD), and patients with late-onset mild cognitive impairment (LMCI). The parameters were inferred from two major AD biomarkers (CSF-Aβ42 and beta-amyloid density) in research subjects from the ADNI database. **A**. Scatter plot of CSF-Aβ42 vs beta-amyloid load for the three groups. Lines represent best fits by Eq. (1) for each group. Vertical dotted lines show the range of the PET signal corresponding to the low amyloid load in panel C. **B**. Heat maps of the posterior probability distributions of the parameters characterizing beta-amyloid turnover in the three groups. **C**. Average CSF-Aβ42 in patients that have a low or high amyloid load. The values calculated using Eq. (1) based on amyloid load with best-fit parameters (solid bars) are not statistically different from the values calculated using CSF-Aβ42 data explicitly (striped bars). The average CSF-Aβ42 in patients with a high amyloid load, regardless of their group, is significantly lower than the average in patients of the corresponding group that have a low load. CSF-Aβ42 is also progressively lower in patients with a more severe clinical condition, regardless of load. However, the average CSF-Aβ42 in subjects with normal cognition and a high load is not different from the average CSF-Aβ42 in patients with AD and a low load.

To elucidate those differences, we fitted the model described in the previous section to the data for each group (see also Methods) and calculated the PDF of the normalized amyloid synthesis and removal, *SYN* and *KF* (see previous section). The lines in Fig. 3A correspond to the most probable values of the parameters in each group, revealing a tendency for the CSF-Aβ42 to be progressively lower with increasing severity of cognitive impairment (consistent with observations by Sturchio et al. [37]). To characterize the statistical significance of the changes in the parameter values, we calculated marginal PDFs of the parameters, shown as heat map plots for each group in Fig. 3B. The boundary between blue and black approximately corresponds to the boundary of the 95% confidence region. The regions appear not to overlap for the NC, LMCI and AD groups. This indicates a statistically significant difference between the estimated parameter values. The difference was independently confirmed by a multi-variate z-test (p<0.05). The estimate of the amyloid removal rate is higher in AD patients compared to NC subjects (see more about the comparison in the next section).

To illustrate that our model accurately predicts average CSF-Aβ42 based on amyloid load, we separated all amyloid-positive subjects in each group into those with low and high amyloid load (20<CL<50 and CL>50, respectively). For each research subject, we used either measured CSF-Aβ42 values or the values calculated based on [PET] value for this subject and the model parameters for the corresponding group. The averages of the values calculated using the model (darker bars in Fig. 3C) were not statistically different from averaged measured CSF-Aβ42 (lighter bars), confirming goodness of fit.

### Patients with early-onset mild cognitive impairment (EMCI) have parameters of Aβ42 turnover similar to those of NC subjects, unlike patients with late-onset mild cognitive impairment (LMCI)

We extended our analysis to include the group of patients with EMCI also available in the ADNI database. Fig. 4A shows 95% confidence regions of the model parameter estimates for NC, EMCI, and LMCI groups. Patients with LMCI and NC subjects represent statistically different groups, whereas confidence regions for the EMCI and NC groups overlap. The latter suggests that EMCI patients may have beta-amyloid turnover parameters that are no different from those in NC subjects.

**Figure 4.**
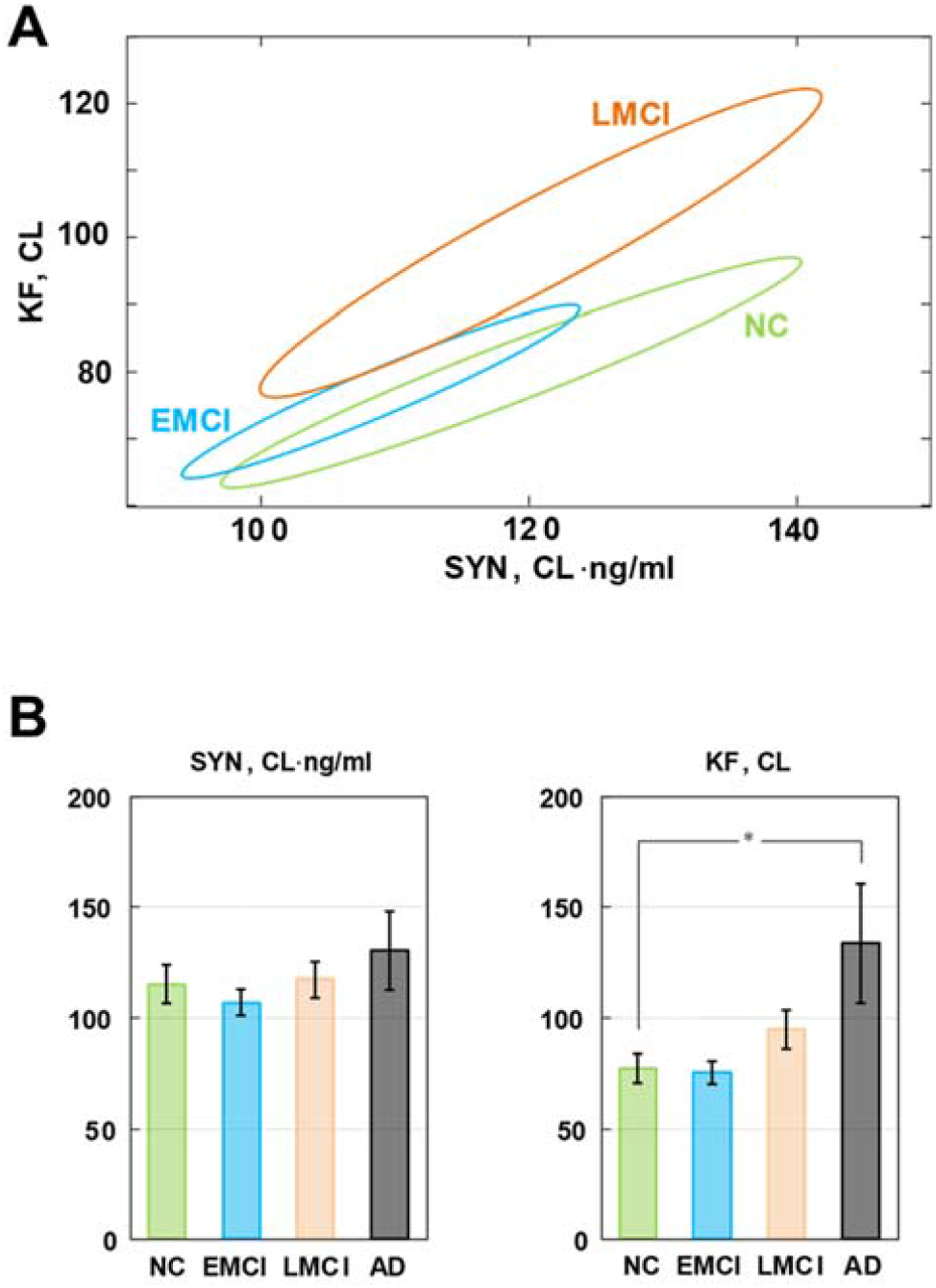
A comparison of beta-amyloid turnover parameters in subjects with normal cognition (NC), patients with either early-onset and late-onset mild cognitive impairment (EMCI and LMCI), and patients with Alzheimer’s disease (AD). **A**. The 95% confidence regions of the parameters characterizing beta-amyloid turnover in the NC, EMCI, and LMCI groups. The confidence regions for the NC and LMCI groups do not overlap, while the confidence regions for the NC and EMCI groups do. **B**. The inferred values of the beta-amyloid synthesis rate (SYN) and the removal rate (KF) for all studied groups. * The values of KF for the NC and AD groups are statistically different (z-test, p<0.05).

To characterize the effect of cognitive impairment severity on individual parameters, we also calculated marginal distributions for *SYN* and *KF*. Fig. 4B shows the results in the form of the mean +/- SD for each parameter in each group. Due to a significant correlation between the parameters (note the elongated shapes of the confidence regions in Figs. 3B and 4A), the only significant difference that could be detected was in the KF value between the NC and AD groups. Based on this, we conclude that AD patients have a significantly higher normalized amyloid removal rate compared to cognitively normal participants, that there is a statistically significant difference between patients with LMCI and NC subjects when both parameters, SYN and KF, are taken into account, and that the EMCI and NC groups do not appear to be statistically different.

### The estimated beta-amyloid cellular uptake is dramatically higher in AD patients compared to NC subjects

Our calculations show that the normalized amyloid removal rate, *KF*, was about 80 (in units of centiloids, CL) in the NC group compared to 140 in the AD group which is a 75% difference. However, our model does not allow for the independent estimation of the components of *KF* - cellular beta-amyloid uptake and removal of beta-amyloid through CSF. As we speculate in the Discussion, the latter component is either not changed in AD patients [12] or is decreased [35]. Therefore, the increase in the amyloid removal rate likely happens exclusively through an increase in cellular beta-amyloid uptake, and hence the increase of uptake in AD patients is likely to exceed 75%.

The estimate of a difference in beta-amyloid cellular uptake between NC and AD participants depends on the distribution of the amyloid removal rate between cellular amyloid uptake and amyloid removal through CSF (see Methods). For the reasons outlined in the Discussion, it is likely that amyloid removal through CSF dominates over the cellular uptake. To examine possible scenarios, we considered cellular uptake to be 50% and 25% of the aggregation-independent removal rate in NC subjects (Fig. 5A). To account for a 75% increase in the total amyloid removal rate in AD patients, beta-amyloid cellular uptake rate should be increased by 2.5 and 4 times, respectively.

**Figure 5.**
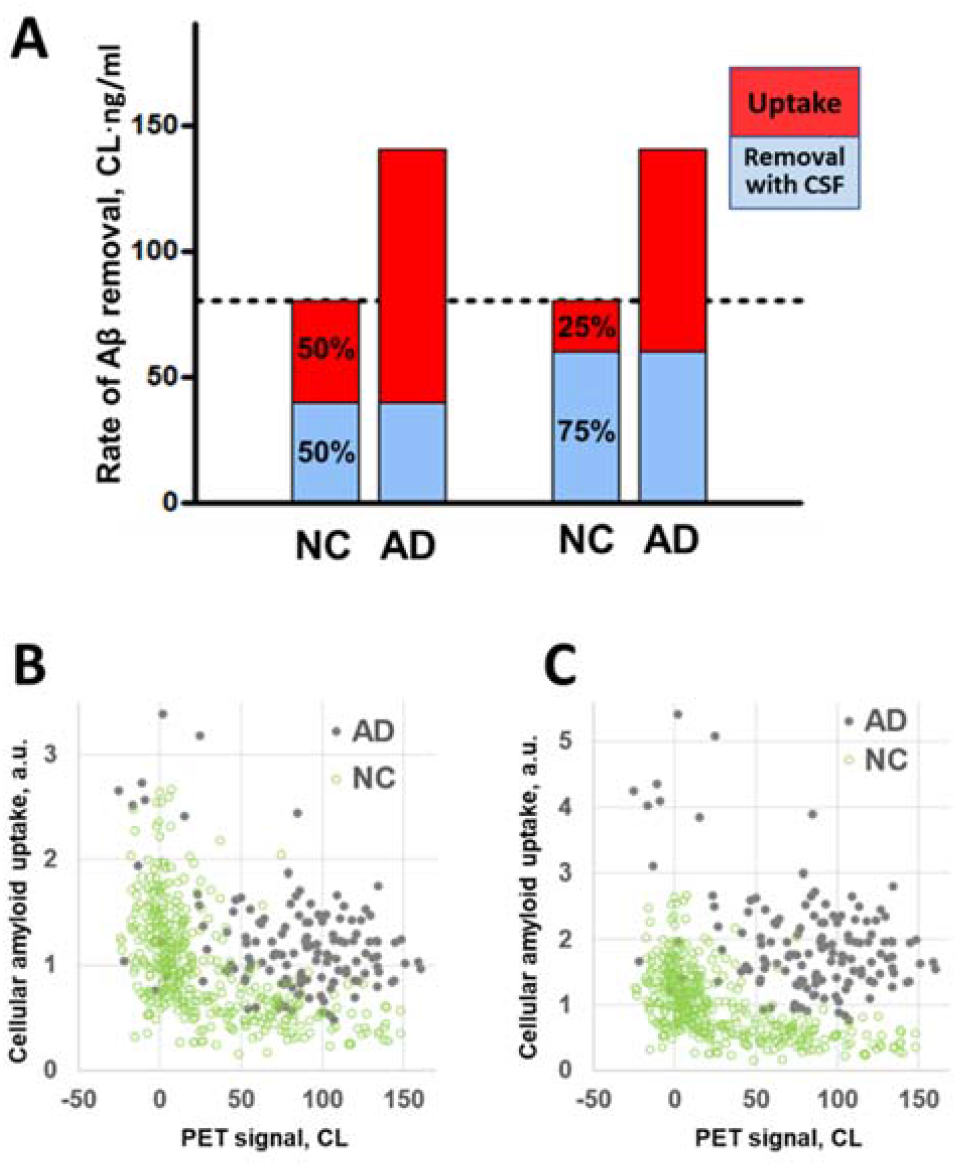
The difference in the aggregation-independent amyloid removal rate between NC participants and AD patients translates into a much greater relative difference in cellular amyloid uptake rate. **A**. The red and blue parts of the bars represent the cellular amyloid uptake rate and the rate of removal through the CSF (the components of the rate of amyloid removal), respectively. If the ratio of the two components is 50/50 in the NC group, the cellular uptake rate is 2.5 times greater in the AD group than in the NC group. If the ratio is 75/25, the difference is 4-fold. **B**. Mass of cellular amyloid uptake (in arbitrary units, a.u.) calculated for individual data points in the NC and AD groups. The rate of removal through the CSF is equal in both the NC and AD groups, and in this scenario is 50% of the amyloid removal rate of the NC group. **C**. Mass of cellular amyloid uptake (in arbitrary units, a.u.) calculated for individual data points in the NC and AD groups. The rate of removal through the CSF is equal in both the NC and AD groups, and in this scenario is 75% of the amyloid removal rate of the NC group.

Using this approach, it is possible to compare the values of the total amyloid uptake - the product of the uptake rate and amyloid concentration in the CSF for individuals in the AD and NC groups from the ADNI cohort (Fig. 5B, C) for the scenarios considering cellular uptake as 50% and 25% of the aggregation-independent removal rate in NC subjects. CSF-Aβ42 is slightly lower in AD patients on average (Fig. 3C), but due to high subject-to-subject variability this effect is hardly seen at the individual level as CSF measurements heavily overlap for any given PET (Fig. 3A). In contrast, the total amyloid uptake appears significantly higher in the AD group (Fig. 5B, C). In case of 4-fold difference in the amyloid cellular uptake rate, the cellular amyloid uptake values for AD patients barely overlap with the total amyloid uptake values for NC subjects regardless of their amyloid load (Fig. 5C).

## DISCUSSION

### PET signal and CSF-Aβ42 are partially independent biomarkers

The brain amyloid load and CSF-Aβ42 are strongly and negatively correlated [10, 11, 21]. It is widely accepted that negative correlation between amyloid plaque density and CSF-Aβ42 occurs because senile plaques create a sink for extracellular Aβ42. In fact, the difference in CSF-Aβ42 between AD patients and NC subjects is in the range of 500 pg/ml [10], while the CSF flow for all subjects is approximately 20 ml/h [35]. The “sink” effect, therefore, should “preserve” approximately 10 ng/h, or close to 100 μg/year. The average extra Aβ42 load of the AD brain compared with a healthy brain is about 5 mg [31] which can indeed accumulate over 50 years. This makes the “sink” theory quite reasonable as an interpretation of the negative correlation between the CSF-Aβ42 and the PET signal.

However, it was shown that these two parameters provide partially independent information [21], as models that include both CSF-Aβ42 and PET measurements have better predictive power for the datasets recorded in different patient cohorts. Specifically, Sturchio et al. [37] demonstrated that in the models adjusted for age, sex, education, APOE4, p-tau levels, and t-tau levels, lower CSF-Aβ42 levels are associated with a higher probability of AD and worse cognitive status, as well as various other indices of the disease. Our analysis confirms that while the biomarkers have strong negative correlation described by a single compartment model, CSF-Aβ42 in fact provides independent clinically valuable information. In cohorts with a similar amyloid load, higher CSF-Aβ42 clearly correlates with a better clinical outcome. CSF-Aβ42 levels above 1000 pg/ml are associated with a low number of AD patients even in the group with high amyloid loads (Fig. 1). Lower CSF-Aβ42 corresponds to a greater increase in the fraction of AD patients if amyloid deposit density is high.

### Cellular amyloid uptake rate may be dramatically higher in AD patients

Sturchio et al. [37] suggested that a high CSF-Aβ42 level plays a critical role in preserving proper brain function. Considering the challenges to this interpretation mentioned in the Introduction, we offer an alternative interpretation by analyzing the characteristic rates of amyloid turnover that we estimated from the same dataset. The model connecting CSF-Aβ42 to the density of amyloid deposits has several parameters which can all potentially be altered by AD. These parameters include the cellular amyloid synthesis rate, the rate of removal through CSF, and the cellular amyloid uptake rate. Due to its biophysical nature, the aggregation rate is considered the same for all populations. Based on Eq. (1), for CSF amyloid concentration to be lower, either the synthesis rate should be lower, or the CSF removal rate should be higher. The suggestion of a lower synthesis rate in AD is not only counterintuitive, but is also not supported by experimental data [23]. With this in mind, the only remaining parameter involved in the turnover of Aβ42 that can explain the difference in CSF-Aβ42 levels between healthy and sick individuals is the amyloid removal rate. Our model predicts a 75% higher amyloid removal rate from interstitial fluid in AD patients compared to cognitively normal individuals. As noted, amyloid removal occurs through both the CSF and cellular uptake, with the removal through the CSF accounting for a greater part of this process.

In the scenarios where cellular uptake is between 50% and 25% of the beta-amyloid removal rate, a 75% increase in the aggregate amyloid removal rate translates into a 2.5 to 4-fold increase in the cellular amyloid uptake rate. Furthermore, amyloid removal through CSF could be decreased in AD patients due to a lower CSF flow [35]. Therefore, to account for the estimated increase in the aggregate amyloid removal rate, the cellular uptake must increase even stronger. We do not have data to estimate the upper boundary of this change, but even the lower boundary of a 2.5-fold increase in the cellular amyloid uptake rate is significant.

According to the model, the increased amyloid removal rate leads to a lower amyloid concentration in the CSF (see Eq. (1)). The absolute amount of amyloid taken by the cells is equal to the product of the cellular uptake rate and the amyloid concentration. However, the reduction in amyloid concentration in the CSF of AD patients compared to the concentration in healthy participants is only about 20-25%, while there is at least a 2.5-fold increase in the cellular amyloid uptake rate (Figs. 3, 4). Therefore, the amount of amyloid taken by the cells can be more than two times greater in AD patients compared to cognitively normal individuals. As we discuss below, this difference can be an etiology-based biomarker of AD.

### Increased cellular amyloid uptake may be one of the key molecular mechanisms defining the age-related progression of Alzheimer’s disease

The progression of Alzheimer’s disease is associated with neuronal death. The severity of the disease correlates with the density of beta-amyloid deposits. Although the aggregated amyloid is not toxic to cells, the synaptic loss and neuritic dystrophy are highest in the anatomical proximity to the senile plaques [27]. On the other hand, it is well-established that soluble beta-amyloid is toxic to neurons both *in vitro* and *in vivo* [16, 33, 38], however the mechanism of its toxicity is still under debate [26, 36].

Recently, we introduced the amyloid degradation toxicity hypothesis that explains known facts about beta-amyloid toxicity and suggests specific pathways leading to cell death [44]. According to the hypothesis, the formation of toxic amyloid products is initiated by the endocytosis of beta-amyloid and the merging of the amyloid-laden endosomes with lysosomes. Intralysosomal digestion produces short fragments of the beta-amyloid peptide, which can create channels in the membranes of lysosomes [24, 41, 42]. These channels are non-selective and can be extremely large [3, 4, 20, 24, 34]. Channel formation is frequently cited as a molecular mechanism underlying AD [2, 6, 14, 15, 18, 19], but it is usually implied that the channels are formed in the plasma membrane, which does not explain many phenomena associated with the disease [45]. Our hypothesis suggests that amyloid membrane channels are formed in lysosomal membranes rather than in plasma membranes. Due to their composition, lysosomal membranes are a perfect target for membrane channel formation by beta-amyloid fragments [41]. The permeabilization of lysosomal membranes in cells exposed to beta-amyloid has been demonstrated by independent laboratories [17, 40]. It readily explains lysosomal/autophagy disfunction, which is considered a major feature of AD [9, 28, 29], as well as intra-cellular ion concentration perturbations induced by exposure to beta-amyloid [43]. Furthermore, lysosomal permeabilization can result in cell death through multiple mechanisms, which we have described previously [44].

An increased cellular uptake rate of beta-amyloid in our model is equivalent to a more intense amyloid endocytosis, which in AD patients results in a greater probability of lysosomal permeabilization and, subsequently, faster cell death.

### AD and EMCI are caused by different pathophysiological mechanisms

Our results suggest that AD and LMCI groups are characterized by an increased cellular amyloid uptake which presumably results in an increased formation of toxic amyloid species and leads to irreversible neuronal damage. In contrast, beta-amyloid turnover in patients with EMCI does not appear different from that in NC subjects. Specifically, there is no tendency towards higher amyloid uptake in EMCI patients (Fig. 4B). Is it possible that beta-amyloid is more toxic in certain patients, even if their amyloid uptake rate is not elevated?

To answer this question, it is necessary to consider how amyloid toxicity develops at the cellular level. Production of peptide fragments is a part of lysosomes’ physiological function, which is to degrade large molecules such as proteins. Unlike short amyloid fragments, full-length Aβ42 is not able to form membrane channels in cell-like structures such as liposomes [41, 42]. Taken together, these facts allow us to hypothesize on the general sequence of intracellular events leading to cell death (Fig. 6). Some beta-amyloid fragments are able to form membrane channels, and some are not [18, 24, 41, 42], so endocytosed beta-amyloid can be degraded by lysosomal proteases into both channel-forming and non-channel-forming fragments (Fig. 6, dichotomy 1). In this context, the channel-forming ability of amyloid fragments is synonymous to their toxicity. Channel-forming fragments such as Aβ25-35, being relatively large peptides, are degraded further (Fig. 6, dichotomy 2). Therefore, toxic amyloid degradation products have a limited life span, and membrane channel formation is most likely a relatively rare event [42]. However, once giant membrane channels are formed in lysosomes, they can leak lysosomal enzymes into the cytoplasm [45] which in turn leads to the activation of necrosis and/or apoptosis.

**Figure 6.**
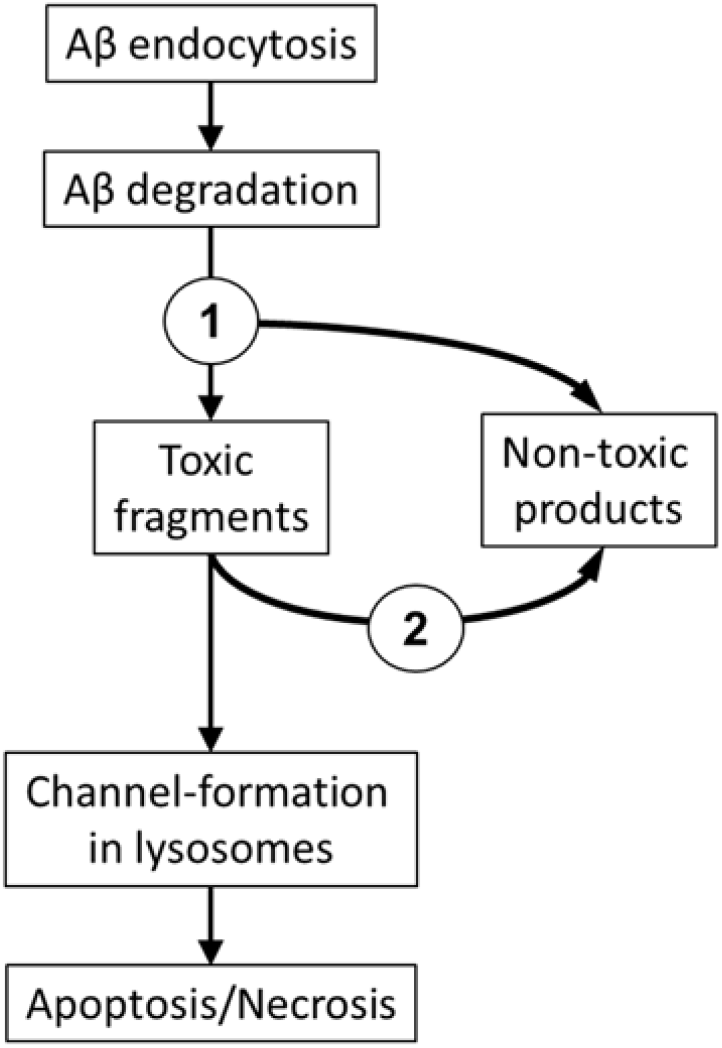
The sequence of events resulting in neuronal death and the progression of Alzheimer’s disease as suggested by the amyloid degradation toxicity hypothesis.

Based on the above, the rate of channel formation depends on the concentration of toxic fragments. This concentration, in turn, depends not only on the concentration of the full-length peptide but also on the balance between the formation and degradation of these fragments. Hypothetically, most toxic amyloid fragments are produced by endoproteases, while their degradation is performed by exoproteases. Therefore, the degradation is mediated by proteases that are not necessarily the same enzymes as the ones producing toxic amyloid fragments. A change in the balance between exo- and endoproteolytic activities of lysosomal proteases would affect the probability of membrane channel formation.

Thus, enzymatic disbalance (an increased rate of toxic fragment production and/or a slower degradation of toxic fragments) can be a mechanism for EMCI that is not associated with an increased cellular uptake rate of beta-amyloid. Alternatively, the concentration of toxic fragments can be increased due to a higher amyloid endocytosis rate, which appears to be the mechanism of AD and LMCI.

### Limitations and future directions

The single compartment model which we used in this study is oversimplified. First, we assumed that CSF-Aβ42 levels can be used as a surrogate for interstitial concentrations of Aβ42. This assumption is based on the correlation of the concentrations of the peptide in these two bioliquids [8], while there is in fact a complex process of amyloid exchange between the interstitial space and the CSF [32]. Second, the model assumes a homogeneous Aβ42 distribution within the interstitial compartment, however, different areas of the brain can have different synthesis and uptake rates of beta-amyloid. Nevertheless, the proposed model allows for the inference of the parameters characterizing beta-amyloid metabolism as an average over the entire brain. These findings add an organ system level into the integrative model of AD pathophysiology which we have proposed previously [44].

The precision of estimates is dependent on the number of observations, so increasing the size of the cohort can demonstrate statistical significance for the differences between the characteristic parameters of the groups. However, using a statistical approach does not allow for the estimation of the parameters of interest in specific patients. Establishing novel biomarkers, which directly characterize the rates of relevant processes such as cellular amyloid uptake and metabolic production of toxic amyloid species, can be a breakthrough for early and/or differential diagnosis of Alzheimer’s disease.

## CONCLUSIONS

Based on this modeling study, we conclude that lower levels of soluble Aβ42, considered independently of the amyloid load, are due to an increased cellular amyloid uptake in the brain. We suggest that the rate of beta-amyloid uptake can be used as a novel pathophysiologically relevant biomarker of Alzheimer’s disease. A differential diagnosis of AD also requires an assessment of lysosomal proteolytic activity that can lead to the production of channel-forming amyloid fragments.

## Data Availability

All data produced in the present study are available upon reasonable request to the authors

http://adni.loni.usc.edu/about/

## Abbreviations

AD: Alzheimer’s disease
MCI: mild cognitive impairment
LMCI: late-onset MCI
EMCI: earlyonset MCI
NC: normal cognition
Aβ: beta-amyloid
Aβ42: Aβ1-42
CSF: cerebrospinal fluid
CSF-Aβ42: concentration of Aβ42 in the CSF

## Acknowledgements

Research reported in this publication was not supported by any external funding.

Data collection and sharing for this project was funded by the Alzheimer’s Disease Neuroimaging Initiative (ADNI) (NIH Grant U01 AG024904) and DOD ADNI (award number W81XWH-12-2-0012). ADNI is funded by the National Institute on Aging, the National Institute of Biomedical Imaging and Bioengineering, and through generous contributions from the following: AbbVie; Alzheimer’s Association; Alzheimer’s Drug Discovery Foundation; Araclon Biotech; BioClinica, Inc.; Biogen; Bristol-Myers Squibb Company; CereSpir, Inc.; Cogstate; Eisai Inc.; Elan Pharmaceuticals, Inc.; Eli Lilly and Company; EuroImmun; F. Hoffmann-La Roche Ltd and its affiliated company Genentech, Inc.; Fujirebio; GE Healthcare; IXICO Ltd.; Janssen Alzheimer Immunotherapy Research & Development, LLC.; Johnson & Johnson Pharmaceutical Research & Development LLC.; Lumosity; Lundbeck; Merck & Co., Inc.; Meso Scale Diagnostics, LLC.; NeuroRx Research; Neurotrack Technologies; Novartis Pharmaceuticals Corporation; Pfizer Inc.; Piramal Imaging; Servier; Takeda Pharmaceutical Company; and Transition Therapeutics. The Canadian Institutes of Health Research is providing funds to support ADNI clinical sites in Canada. Private sector contributions are facilitated by the Foundation for the National Institutes of Health (www.fnih.org). The grantee organization is the Northern California Institute for Research and Education, and the study is coordinated by the Alzheimer’s Therapeutic Research Institute at the University of Southern California. ADNI data are disseminated by the Laboratory for Neuro Imaging at the University of Southern California.

The authors thank Nikita Zaretsky for his editorial help.

